# Epidemiology and Characteristics of MPXV Clade I cases in the WHO European Region, 2024-2025

**DOI:** 10.64898/2025.12.17.25342473

**Authors:** Ioannis Karagiannis, Jeffrey Pires, Pana Akhmetniyaz, Olivier le Polain de Waroux, Bart Hoorelbeke, Bertrand Gagnière, Klaus Jansen, Chrysa Tsiara, Derval Igoe, Maria Rosaria Campitiello, Catharina van Ewijk, Vítor Cabral Veríssimo, Laura Santos Larrégola, Erik Sturegård, Céline Gardiol, Taliha Karakök, Gareth J. Hughes, Marc-Alain Widdowson

## Abstract

**BACKGROUND:** A public health emergency of international concern (PHEIC) was declared in August 2024 following a sharp increase in mpox cases linked to the emergence of monkeypox virus (MPXV) clade Ib.

**AIM:** To characterise imported and locally acquired mpox clade I cases reported in the WHO European Region since August 2024, and to assess transmission patterns and response measures.

**METHODS:** Between 14 August 2024 and 23 November 2025, we collected key information on MPXV clade I infections reported by member states of the WHO European Region through the International Health Regulations. We contacted reporting countries to collect age, sex, travel history, most likely route of transmission, clinical course, contacts and type of exposure, and implemented control and prevention measures.

**RESULTS:** Eighty-two cases of MPXV clade I infection were reported; forty-five were imported and 37 were infections acquired in reporting member states. Seventy-nine were typed as clade Ib and two as clade Ia. Most imported cases reported heterosexual or close physical contact as possible exposure. Secondary transmission occurred in six households. One healthcare worker was infected. Since October 2025, 17 autochthonous cases in men were most likely infected through sex with other men.

**CONCLUSIONS:** Imported cases of MPXV clade I infection were associated with limited household transmission. The increase in autochthonous infections among men with recent sexual contact with other men suggests undetected spread in Europe, that may become sustained. Continued surveillance, case and contact investigation are needed to understand MPXV clade I epidemiology and drivers of MPXV clade I transmission in Europe.

## Introduction

Two major genetic clades of monkeypox virus (MPXV) have been described: clade I, historically associated with central Africa, and clade II, historically associated with west Africa [1-2]. Before the recent multi-country outbreaks, cases outside Africa were rare and associated with little or no onward transmission [2-3].

In 2020, the largest recorded outbreak of mpox due to MPXV clade I occurred in the Democratic Republic of the Congo [4] and in 2022, a global mpox epidemic caused by MPXV clade II prompted the World Health Organization (WHO) Director-General to declare a Public Health Emergency of International Concern (PHEIC) [5]. The monthly number of reported global cases peaked at approximately 30,000 cases in August 2022 [6], followed by a decline in cases globally and in Europe [7] and in May 2023 the PHEIC was lifted [8]. Since September 2023, the Democratic Republic of the Congo has seen a sharp increase in the reported number of mpox cases, linked to both the emergence of a distinct new strain, MPXV clade Ib, and a rise in cases caused by infection with previously circulating MPXV clade Ia [9, 10]. Earlier evidence, including a 2022 meta-analysis, suggested that MPXV clade I may be associated with higher case-fatality than clade II [9, 11]. The newly identified clade Ib rapidly spread to neighbouring African countries and elsewhere. Initial reports from Democratic Republic of the Congo indicated sustained person-to-person transmission, including through heterosexual sex. For this reason, WHO declared mpox a PHEIC for a second time on 14 August 2024 [12], which was lifted in September 2025 [13]. The first case of MPXV clade Ib infection outside of Africa was reported by Sweden on the next day [14, 15]. Between January 2024 and 7 November 2025, 17 countries in the WHO African Region and 26 countries in all other regions have reported MPXV clade Ib cases [6], with the European region reporting most MPXV clade I cases. Evidence from imported cases [16, 17] and outbreaks in Africa [10, 18] indicates that MPXV clade I transmission may occur both within and outside sexual networks, including through skin-to-skin contact [19].

Rapid investigation of MPXV clade I cases and timely public health response are essential to prevent further spread, provide key data on the epidemiology of MPXV clade I and inform risk assessments for Europe and comparable settings worldwide. In this paper, we describe MPXV clade I cases reported in the WHO European Region since August 2024 to gain a better understanding of the characteristics and epidemiology of MPXV clade I cases in Europe.

## Methods

### Data collection

The WHO Office for the European Region collected information on MPXV clade I cases reported in line with WHO standing recommendations for mpox [20] through International Health Regulations (IHR) channels [21] between 14 August 2024 and 23 November 2025. Reporting MS were contacted either by email or teleconferences together with relevant colleagues from the European Centre for Disease Prevention and Control (ECDC) using a standard set of questions to collect information on age, sex, travel history, likely route of transmission, clinical course and hospitalisation, contacts, and implemented control and prevention measures.

### Case definitions

Confirmed cases were defined as persons with or without symptoms, with laboratory confirmation of MPXV clade I by molecular detection in any clinical specimen, including lesion swabs and upper respiratory tract specimens. Probable cases were defined as persons with symptoms compatible with mpox who were epidemiologically linked to a confirmed case, but without laboratory confirmation of MPXV clade I.

Cases were classified as imported if their personal history indicated that they had contracted infection outside the reporting MS in the 21 days preceding symptom onset during national case investigation activities or, otherwise, as autochthonous. Clusters of cases were defined as individual cases who were epidemiologically linked.

### Laboratory confirmation

Laboratory confirmation and clade classification were performed by national or subnational laboratories in the reporting Member States (MS) as part of routine public health investigation. Cases included in this analysis had molecular confirmation of MPXV clade I by PCR-based testing performed on clinical specimens. Subclade assignment (Ia/Ib) was based on national laboratory typing and, where available, sequencing or clade-specific molecular characterisation. Laboratory approaches were not standardised across countries. Published descriptions of microbiological methods for selected European cases are available elsewhere [14,17,22-24].

### Statistical analysis

We described cases by age, sex, travel history, possible exposures, most likely route of transmission, hospitalisation and clinical course, care-seeking behaviour, diagnosis delays, HIV status or other clinical comorbidities that cause immunosuppression, prior vaccination for mpox, number of contacts, and information on implemented control and preventive measures. Continuous variables across case groups were compared using the Kruskal-Wallis test using Stata v19.5. Percentages have been rounded to the nearest decimal.

## Results

As of 23 November 2025, 13 of the 53 MS reported one probable and 81 confirmed cases of MPXV clade I infection to the WHO Regional Office for Europe; one case reported by Türkiye and one reported by the Republic of Ireland were typed as clade Ia, and the remaining 79 were typed as MPXV clade Ib; the MPXV clade Ia case reported by the Republic of Ireland has been described elsewhere [22]. The other, a probable case reported by the Republic of Ireland, was imported and had close contact with a subsequently confirmed case of MPXV clade Ib infection. One case with travel history to Asia initially notified as MPXV clade Ib was later found to be a recombinant virus containing elements of both clade Ib and IIb [23]. Of the 82 cases, 45 (54.9%) were imported and 37 (45.1%) were autochthonous. Likely countries of infection included Uganda (N=11), Democratic Republic of the Congo (N=6), Rwanda (N=5), the United Republic of Tanzania (N=5), Kenya (N=3), the United Arab Emirates (N=3), Thailand (N=2), the Netherlands (N=2), Malaysia (N=1), Pakistan (N=1), Viet Nam (N=1), Angola (N=1), “East Africa” (N=1), and “Central Africa” (N=1). One case had travelled to Rwanda, Uganda, and United Republic of Tanzania within 21 days before symptom onset, and one case had travelled to both Egypt and United Arab Emirates, but case history suggests transmission most likely occurred in the latter. Of the 37 cases infected in Europe, 17 were secondary cases of one of seven imported cases or an already known autochthonous case, and a further 20 cases were not related to any reported case (Figure 1).

**FIGURE 1:**
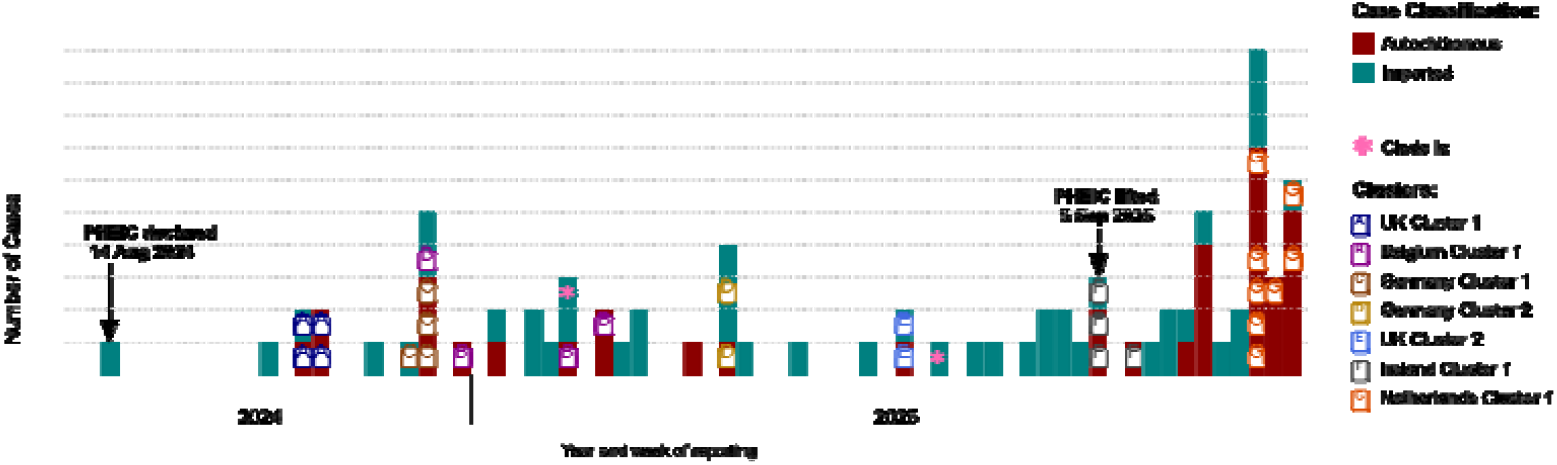
Reported MPXV clade I cases and clusters, 14 August 2024 – 23 November 2025, WHO Regional Office for Europe (N=82).

Of the 45 imported cases, 43 were male and two were female. Age was known for 38 cases, while only the 10-year age group was known for another six; the median age was 36.5 years (range 18-64 years). The most likely route of infection was assessed as sexual contact for 26 of 45 imported cases, other intimate physical contact (e.g. massage, sauna services) for five and unknown for the remaining 14 cases (Table 1). Among the 25 imported men reported to have acquired infection sexually, the sex of the most likely infecting partner was known for 17; in 12 cases the partner was female and in five the partner was male. One additional imported case with sexual exposure was female.

**TABLE 1:** Reported probable mode of transmission of MPXV clade I cases, WHO Member States, 14 August 2024 – 23 November 2025 (N=82)

| Probable mode of transmission | Imported cases (N=45) | Autochthonous cases (N=37) |  | All cases (N=82) |
| --- | --- | --- | --- | --- |
|  |  | Household cases (N=13) | Other cases (N=24) |  |
| Sexual contact | 26 (57.8%) | 4 (30.8%) | 19 (79.2%) | 49 (59.8%) |
| Other contact in household | 0 (0%) | 9 (69.2%) | 0 (0%) | 9 (11.0%) |
| Close physical contact (e.g. sauna, spa, massage) | 5 (11.1%) | 0 (0%) | 0 (0%) | 5 (6.1%) |
| Healthcare setting | 0 (0%) | 0 (0%) | 1 (4.2%) | 1 (1.2%) |
| Unknown | 14 (31.1%) | 0 (0%) | 4 (16.7%)<sup>a</sup> | 18 (22.0%) |

<sup>a</sup> One of these cases reported contact with family members who had travelled to an outbreak-affected country in Central Africa.

Information on hospitalisation was available for 79 cases, of whom 18 (22.8%) were hospitalised; four, among whom one child, were admitted for clinical reasons and 13 for isolation or other non-mpox-related reasons. The reason for hospitalisation was not available for one hospitalised case. Prior vaccination status was available for 51 cases, among whom four had been vaccinated before symptom onset. The exact vaccine product was not available. HIV status was known for 44 cases, among whom six were living with HIV. Information on immunocompromising conditions other than HIV was available for 36 cases, none of whom were reported to be immunocompromised. All cases have recovered.

Of the 37 autochthonous cases, 25 were male and 10 were female; sex information was unavailable for two cases. Thirteen were household contacts of imported cases.

Of these 13 secondary cases in households, seven were below the age of 15 years (three were aged 1-4 years, two were aged 5-9 years, and the remaining two were 10-14 years old). Four of the 13 household cases were partners of index cases (one of whom was a probable case) and were reported to have contracted mpox sexually from their partners; the remaining nine likely acquired infection through household contact. One of the cases infected in Europe was a healthcare worker who had cared for a hospitalised case before mpox was included in the differential diagnosis. Standard IPC precautions appropriate to the initial working diagnosis were used, including gown/apron, gloves and a surgical mask; no breach of PPE or needlestick injury was reported. Contact and airborne-based precautions were introduced after mpox was suspected (Table 1) [24].

Of the 24 autochthonous cases with no clear exposure to a reported case, one had no travel history but reported contact with her male partner and another family member who had both returned from a central African country in the previous 21 days; neither of them had reported symptoms compatible with MPXV infection. The exact source of infection for the other cases remains unknown (Table 1). Among 44 men who contracted mpox sexually, information on the sex of the most likely infecting partner was available for 40 of them; among these, 21 reported having most likely acquired the infection from other men most likely in the Netherlands (N=11), Spain (N=8), Malaysia (N=1), and United Arab Emirates (N=1). No men who acquired MPXV clade Ib through sexual contact with other men were reported to WHO before October 2025. Six of the 11 cases reported by the Netherlands and one reported by Belgium reported having attended the same sex venue in Amsterdam. Of these six cases reported by the Netherlands, four attended the venue on the same date, and the remaining two on two different dates; according to the national investigation, all six had attended within one incubation period prior to symptom onset, suggesting that they most likely acquired infection there.

Of the 45 imported cases, information on whether the individual was returning to a household with other residents was available for 34; of these, 25 returned to households with other residents. Transmission was identified in six of these 25 households. Within these six households, 13 additional cases occurred among 33 household contacts (overall attack rate: 39.4%, range 0-100%). Two household cases were determined to be possible second-generation transmission events; in one household, the case had symptom onset in January 2025 following two cases with symptom onset in the same household in December 2024, although transmission through fomites or an unidentified fourth household case could not be excluded. On another occasion, a probable imported case infected his wife sexually (attack rate: 12.5%, 1/8 susceptible household contacts). The wife then likely infected one of the remaining seven susceptible household members through close physical contact (secondary attack rate: 14.3%, 1/7). Of 37 imported cases with information available, 15 travelled to Europe by air while symptomatic or with rash and, in at least one case, with an uncovered rash on their arms.

MS reported that contacts of mpox cases were identified and risk classified in line with WHO [25] or national guidance. However, follow-up of travel-related contacts was not implemented consistently across MS; for one imported case, national health authorities asked the airline to contact the co-passengers who had been exposed to the case on a 1 hour flight.Information on the number of contacts in travel and school settings was sparse, and no secondary case was reported among travel or school contacts.

Where information was available (N=68), subclade assignment was reported at a median of 9.5 days after symptom onset (-1 to 74 days); two patients were diagnosed before symptom onset due to active follow-up following household exposure. In these cases, MPXV was detected by PCR from upper respiratory tract specimens and, where lesions subsequently appeared, lesion specimens. However, specimen source and testing approaches were not systematically available for all contacts across MS.

Contacts of already reported cases sought or received care quicker and were diagnosed sooner than imported cases or those autochthonous cases with no reported exposure to a known case (Table 2); nonetheless, contacts of known cases still took a median of 4 days after symptoms onset to seek care.

**TABLE 2:**
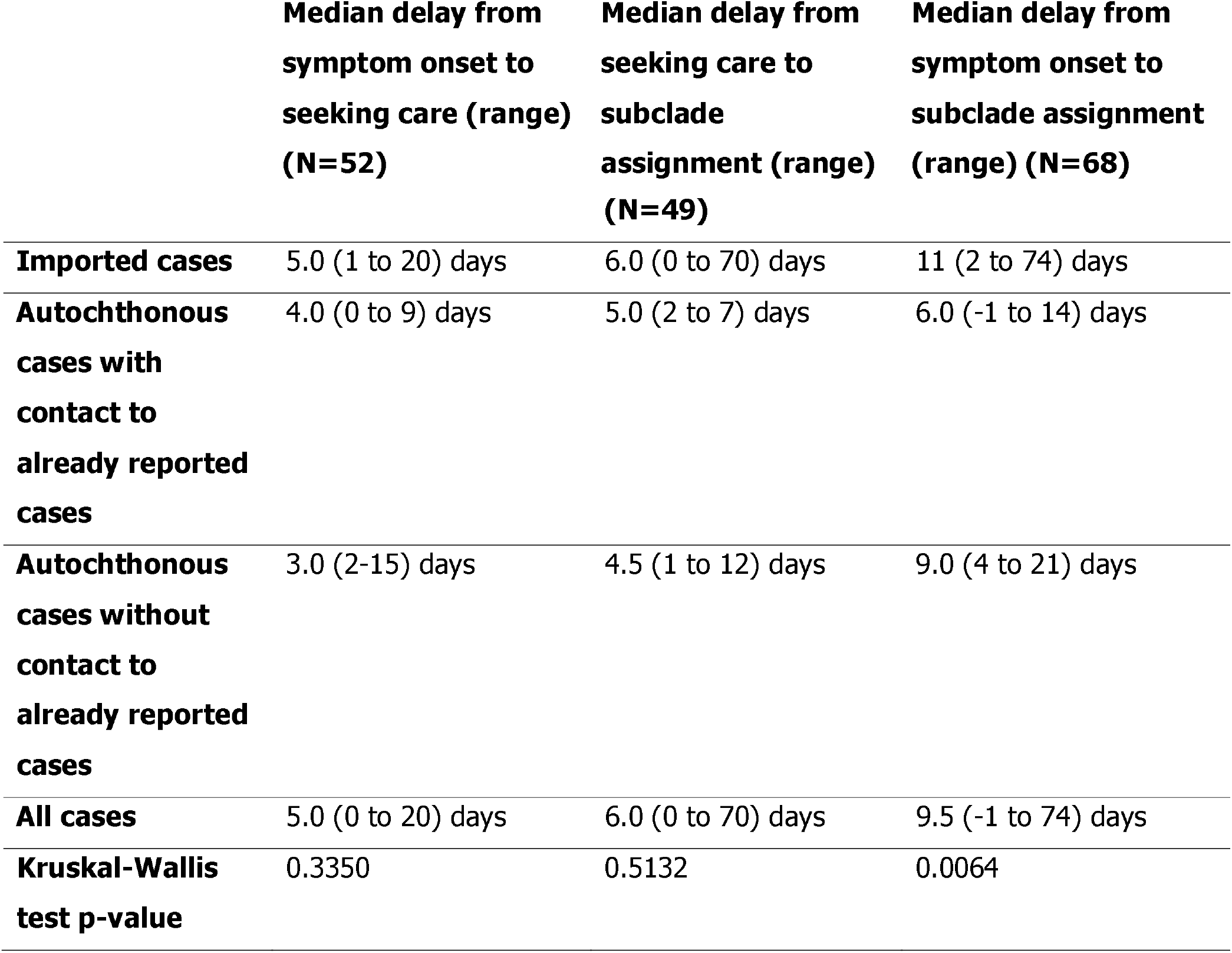
Time to seeking care and to diagnosis of reported clade I MPXV cases, WHO EURO member states, 14 August 2024 – 23 November 2025.

## Discussion

Since the declaration of a PHEIC in August 2024 and until 23 November 2025, 82 confirmed MPXV clade I cases have been reported in 13 member states of the WHO European Region. Until October 2025, most cases were imported, with limited secondary and possibly tertiary household transmission involving partners and children. Since October 2025, an increasing number of cases without links to travel have been reported, especially among men who have sex with men, suggesting circulation of the virus in Europe [26, 27]. Sexual transmission of MPXV clade Ib, especially in MSM networks, appears to have become an important route of transmission in Europe in recent months. Experience from MPXV clade IIb suggests that the spillover risk for the general population remains limited [7].

We describe here also two cases of MPXV clade Ia infection, both with recent travel to the Democratic Republic of the Congo. One of the sequences contained a mutational signature widely reported for MPXV clade IIb virus genomes during the 2022-2023 global outbreak. The presence of this host-driven mutagenesis pattern in a MPXV clade Ia virus suggests substantial person-to-person transmission [28].

Follow-up of flight passengers was implemented inconsistently across MS, which limited assessment of transmission risk during air travel. No secondary cases were identified among reported travel contacts in available data, in line with published evidence suggesting a very low risk of mpox transmission during air travel for MPXV clade IIb [29]. Disentangling transmission routes in household settings was complicated by the sensitive nature of the infection and the multiple possibilities of contact and fomite use. This underlines the importance of reinforcing existing WHO recommendations for household contacts [25]. The healthcare-associated case occurred before mpox was suspected clinically, when the patient was managed using precautions appropriate to an alternative working diagnosis rather than mpox-specific precautions [24]. This highlights the importance of considering mpox early in the differential diagnosis of compatible presentations in healthcare settings.

MPXV clade I infections have been previously considered more severe than clade II infections [9, 30]. However, our data showed that reported cases in the European Region were much less severe than those reported in African countries; no deaths were observed, and among the 79 cases for whom hospitalisation status was available, only four were hospitalised for clinical reasons, including one child. This is consistent with more recent data indicating that, in areas where MPXV clade Ib has become endemic, CFRs have been reported to be below 1% [9]. This may reflect not only differences in the affected populations and transmission patterns, but also earlier health care seeking and access to supportive care. Interpretation of underlying clinical risk factors was limited by incomplete data; among cases with available information, six were reported to be living with HIV, none were reported to be immunocompromised other than HIV, and four had received mpox vaccination before symptom onset.

Since 2023, the outbreak of MPXV clade I in the Democratic Republic of the Congo has affected younger age groups, including children [10, 18]. In our series, sexual contact was the most likely route of transmission for 26 of 45 imported cases. Among the 25 men who were judged to have acquired the infection sexually overseas, the sex of the most likely infecting partner was known for 17, and in 12 of these cases the partner was a woman. Of the 18 locally acquired MPXV clade Ib cases in Europe, seven were among children under 15 years, whereas since 2022, only 38 locally acquired clade II cases in this age group, despite more than 33,000 cases of clade IIb in MSM [6]. This may reflect differences between the predominantly non-sexual household transmission observed following imported MPXV clade I cases, including infections among children, and the ongoing sexual transmission of clade IIb mpox in MSM networks in the WHO European Region. More data is needed to see whether these differences reflect contact patterns, virologic factors, or both.

Although active follow-up of contacts supported the identification of secondary cases in Europe, healthcare-seeking and diagnostic delays, especially among index cases, may have contributed to onward transmission. Delays were substantial in our case series, with a median of 6 days from seeking care to virus subclade assignment and up to 70 days among imported cases. Clinicians in primary and acute care should consider mpox in patients with compatible symptoms regardless of travel history, and request PCR with clade-specific typing at initial testing, given autochthonous MPXV clade Ib circulation in Europe. The 4-day gap between symptom onset and seeking care among contacts of known cases was notable despite active follow-up.

The outbreak in Central and East Africa, albeit declining, is ongoing despite control efforts, including vaccination. While MPXV clade I continues to circulate in these regions, further importations into the European Region are likely. In Europe, the epidemiology of MPXV clade I is changing. Early in the observation period, most cases were imported, often following heterosexual exposure in outbreak-affected countries, with only limited onward transmission, mainly in household settings. More recently, however, an increasing number of autochthonous cases have been reported among MSM, often without clear epidemiological links to known imported cases, suggesting a shift from isolated importation events towards circulation within MSM sexual networks such as for clade IIb. The clustering of cases linked to a sex venue in Amsterdam further supports this interpretation. Similar locally acquired MPXV clade Ib transmission among MSM and their networks was also reported in the United States around the same period [31]. Uncertainties remain regarding the extent of silent importations, age-specific risk, secondary attack rates, and vaccine-derived protection. MPXV clade II transmission continues in Europe at low levels of 100-300 cases a month, mainly in MSM sexual networks [6, 7]. An immediate priority in Europe remains to reduce the risk of sustained MPXV clade I transmission through early detection, interruption of transmission, and protection of groups at higher risk, in line with the WHO European Region goal of mpox elimination. As more recent MPXV clade Ib spread in Europe has been largely driven by MSM sexual networks rather than isolated importations, understanding current immunity levels among high-risk groups is important. Published estimates suggest that mpox vaccine coverage among MSM in Europe is heterogeneous and incomplete, with ECDC citing EMIS-2024 data showing two-dose self-reported coverage ranging from 0.5% to 37.9% across EU/EEA countries, which supports continued emphasis on vaccination in higher-risk groups [32]. Other modelling analyses also point to maintenance of high-risk behaviours among MSM that should be tackled through awareness campaigns [33]. Although the PHEIC was lifted in September 2025, WHO Standing Recommendations [20] remain important for countries to mitigate the establishment of MPXV clade I transmission.

Continued global collaboration on genomic surveillance with subtyping, early case identification, early diagnosis and systematic contact tracing, including timely cross-border information sharing, remain essential to reduce the risk of sustained MPXV clade I transmission in Europe and to understand virological evolution [7, 25]. Strengthening diagnostic capacity, including subtyping, is critical to detect and monitor emerging variants and guide appropriate public health action [28, 34]; the recombinant virus reported by the United Kingdom would not have been identified without routine genomic surveillance [23]. Given the observed cases among a healthcare worker and within an MSM network, countries in Europe should ensure robust infection prevention control in healthcare facilities and sustain engagement with MSM communities.

Finally, renewed efforts should also ensure that vaccination, clear information on infection risk, and behavioural measures to reduce it reach affected groups. These include reducing the number of partners and safer sex practices, especially for individuals who may play a disproportionately high role in transmission within MSM networks.

This analysis has several limitations. We relied on case-based information reported to WHO by MS, and data completeness and detail varied between countries. As WHO works primarily with nationally reported data, some variables were not consistently available or could not be shared in sufficient detail. Laboratory approaches, sample-taking and contact follow-up practices were not standardised across the Region, and countries differed in MPXV testing algorithms, specimen types, and access to clade-specific typing or sequencing. Exposure histories were based on case interviews, and may therefore be affected by recall and social desirability bias, particularly for sensitive exposures. Differences in case detection, contact tracing and follow-up may also have influenced the reported distribution of transmission routes and clusters. Finally, undiagnosed mild or asymptomatic infections and unrecognised transmission chains are likely to have resulted in under-ascertainment.

## Conclusions

Most early MPXV clade I cases reported in the WHO European Region were imported and resulted in limited onward transmission, mainly in household settings. More recently, autochthonous cases among men exposed through sex with other men suggest changing transmission patterns and spread within MSM sexual networks in Europe. Severe clinical outcomes were uncommon in this case series, with no deaths observed and few hospitalisations for clinical reasons. Continued surveillance, early diagnosis, subclade-specific testing, case investigation and contact tracing remain essential to reduce the risk of further spread and possible establishment of MPXV clade I in Europe. Targeted vaccination, risk communication and engagement with affected communities should remain central in the public health response.

## Data Availability

All data produced in the present study are available upon reasonable request to the authors.

## Acknowledgments

We wish to thank Xanthi Andrianou and Gianfranco Spiteri from the European Centre for Disease Prevention and Control for critically reviewing this paper, as well as all colleagues in Europe working on mpox response at the national and subnational level for their continued contributions to surveillance.

## Notes

### Competing Interest Statement

The authors have declared no competing interest.

### Funding Statement

This study did not receive any funding.

### Author Declarations

All data was deidentified before analysis.

### Summary of Updates

Extensive updates based on reviewers' and editorial comments from the Eurosurveillance journal in collaboration with all co-authors.

